# A k-mer based transcriptomics analysis for NPM1-mutated AML

**DOI:** 10.1101/2023.01.31.23285241

**Authors:** Raíssa Silva, Cédric Riedel, Benoit Guibert, Florence Ruffle, Anthony Boureux, Thérèse Commes

**Author notes:** Equal supervisor (Anthony Boureux, Thérèse Commes).

## Abstract

**Motivation:** Acute Myeloid Leukemia is a highly heterogeneous disease. Although current classifications are well-known and widely adopted, many patients experience drug resistance and disease relapse. New biomarkers are needed to make classifications more reliable and propose personalized treatment.

**Results:** We performed tests on a large scale in 3 AML cohorts, 1112 RNAseq samples. The accuracy to distinguish NPM1 mutant and non-mutant patients using machine learning models achieved more than 95% in three different scenarios. Using our approach, we found already described genes associated with NPM1 mutations and new genes to be investigated. Furthermore, we provide a new view to search for signatures/biomarkers and explore diagnosis/prognosis, at the k-mer level.

**Availability:** Code available at https://github.com/railorena/npm1aml and https://osf.io/4s9tc/. The cohorts used in this article were authorized for use.

## 1 Introduction

Acute Myeloid Leukemia (AML), the most common acute leukemia among adults, is a hematopoietic disorder characterized by neoplastic proliferation of myeloid-lineage cells. Most patients are not cured with current therapies, and despite targeted therapeutic proposals, drug resistance and disease relapse remain persistent problems. AML is also a complex disease with a diversity of cell phenotypes. Several classifications are used to improve clinical outcomes, including French-American-British (FAB) (Bennett *et al*., 1976) and European LeukemiaNet (ELN) (Döhner *et al*., 2017). Despite that, new biomarkers are required for more confident classification and to refine treatment follow-up.

Exploration of RNAseq data provides a deep investigation potential to identify a large diversity of biomarkers and to target key molecular mechanisms that drive the AML pathogenesis and progression, thus, offering potential benefits for diagnosis and prognosis applications. However, RNAseq data can produce large and complex amounts of data, often not human-understandable. Methods such as Machine Learning (ML) have been supporting large-scale analysis of RNAseq data for precision medicine (MacEachern and Forkert, 2021).

In the last years, ML has been largely used in the field of human health for diagnosis and prognosis of diseases, including AML context (Eckardt *et al*., 2020). The use of ML to guide analyzes on large amounts of data offers us a way to interrogate specific parts of RNA, at the k-mer level. K-mers are k-length substrings extracted from a raw sequence file and are revolutionizing large-scale RNAseq data analysis by allowing reference-free queries (Marchet *et al*., 2021). The investigation of k-mers can reveal alterations in certain parts of the gene and links with other genes in a deep-seated way, providing a new view of the investigated scenarios. Also, one of the advantages of using k-mers is to account for the vast diversity of transcriptional events such as splicing events, alternative polyadenylation, intron retention, or mutations making the analysis a reference-free approach.

In this context, we investigated gene expression using k-mers and Machine Learning methods to yield a better understanding of prognosis classification and key mechanisms of AML pathogenesis. To apply a reference-free method in the context of AML, we started looking into NPM1 mutation, one of the most common mutations in this pathology, to define distinct patient groups and to find linked genes by searching at the k-mer level from public AML RNAseq cohorts. Furthermore, our approach provides support for indexing k-mers which constitutes an interesting solution for interrogating “Omics data” instead of extracting the presence/absence or absolute count of k-mers directly from the raw data.

## 2 Materials and methods

In this section, we present how we designed the analysis to identify the genes with different expressions in mutated and non-mutated NPM1 AML patients. For that, we applied a k-mer based approach and machine learning classification in different AML cohorts to select the k-mers, and consequently the genes, that distinguish the NPM1 mutation condition.

### 2.1 Cohorts

We analyzed three RNAseq AML cohorts. Beat-AML (Tyner *et al*., 2018), Leucegene (BCLQ, 2019), and Beat-AML2 (Bottomly *et al*., 2022) cohorts with 462, 437, and 213 samples (phs001657.v1.p1, GSE49642, and phs001657.v2.p1 accession IDs), respectively. From these cohorts, the analyzed samples are Bone Marrow and Peripheral Blood sample types. For each sample, we have information about the presence or absence of NPM1 mutation that allows us to investigate the difference between these two conditions and provide a selection of k-mers. Table 1 presents an overview of information from these AML cohorts.

**Table 1.** Overview of AML cohorts. Number of NPM1 mutated and non-mutated patients and samples types.

|  | Mutated | Non-Mutated | Total |
| --- | --- | --- | --- |
| **Beat-AML** | **112** | **350** | **462** |
| Bone Marrow | 60 | 183 | 243 |
| Peripheral Blood | 52 | 167 | 219 |
| **Leucegene** | **139** | **297** | **436** |
| Bone Marrow | 72 | 155 | 227 |
| Peripheral Blood | 67 | 142 | 209 |
| **Beat-AML 2** | **58** | **148** | **206** |
| Bone Marrow | 35 | 83 | 118 |
| Peripheral Blood | 23 | 65 | 88 |

Additionally, we used five wild-type cohorts, totalizing 132 healthy samples, which in this paper, we treat as a single cohort, the healthy cohort that includes CD34+ cells (37 samples), purified monocytes (50 samples) and mononuclear cells (45 samples). We sought to understand the behavior of the selected k-mers in healthy samples. For that, we searched the k-mers expression in the healthy cohort, considering that the healthy cohort does not have an NPM1 mutation. We interpreted that the k-mers expression needs, at least, to be close to zero to have a link to AML conditions. More information about the five wild-type cohorts can be found in the supplementary material.

To complete the analysis, we analysed at the leukemia cell level. We used 34 RNAseq samples from mature and immature blasts cells obtained from 17 AML patients (https://www.ncbi.nlm.nih.gov/bioproject/PRJEB54896 accession ID). We analyze the k-mers expression to have a closer understanding of the genes. For this task, we searched the selected k-mers from the 3 AML cohorts.

### 2.2 Generating and selecting the k-mers

We checked the quality of the raw downloaded data using fastQC version 0.11.9 (Andrews *et al*., 2010) and MultiQC version 1.9 (Ewels *et al*., 2016). As a complementary quality control, we checked sequencing protocol information and contamination with KmerExplor (Riquier *et al*., 2021)).

Then, we used Kmtricks to build a k-mer count matrix for each cohort, totaling five matrices (three AML, one healthy, and one mature and immature blasts cells). Kmtricks (Lemane *et al*., 2022) is a tool to count k-mers efficiently in large datasets and produce a k-mer count matrix across multiple samples. For example, the Beat-AML cohort has 7 terabytes of fastq files, Kmtricks reduced to a k-mer count compressed matrix of 78 gigabytes. To generate the matrices, in the AML cohorts, we considered that the k-mer needs to have a minimum abundance of 4 (which means, the k-mer has to be found at least 4 times in the sample) and be present in at least 5% of the cohort samples to be counted. In the healthy and mature and immature blasts cells cohorts, we only considered that the minimum abundance was 2. The k-mer size applied was the tool default, size 31nt. The parameters used in Kmtricks for each matrix can be found in the supplementary material.

After building the matrices, we implemented a C++ code to select the k-mers with significant differences between NPM1 mutated and non-mutated samples, based on the coefficient of variation of these two conditions. Due to the k-mers expression variation across the cohorts, for each AML cohort we selected manually the coefficient of variation based on the gene investigated. For that, we analyzed the k-mers coefficient of variation of NPM1 across the AML cohorts. We designed all the specific and unique k-mers of the NPM1 gene with Kmerator (Riquier *et al*., 2021) and counted their occurrence in the 3 AML cohorts with a request from the TranSipedia website (https://transipedia.montp.inserm.fr/). From the k-mers counts, we computed the k-mer coefficient of variation across the entire cohort. Figure 1 shows the coefficient of variation of the k-mers for each cohort. Leucegene and Beat-AML 2 showed less variation, thus, we define the coefficient of variation threshold as 0.98, for Beat-AML we define a 0.95 threshold.

**Fig. 1:**
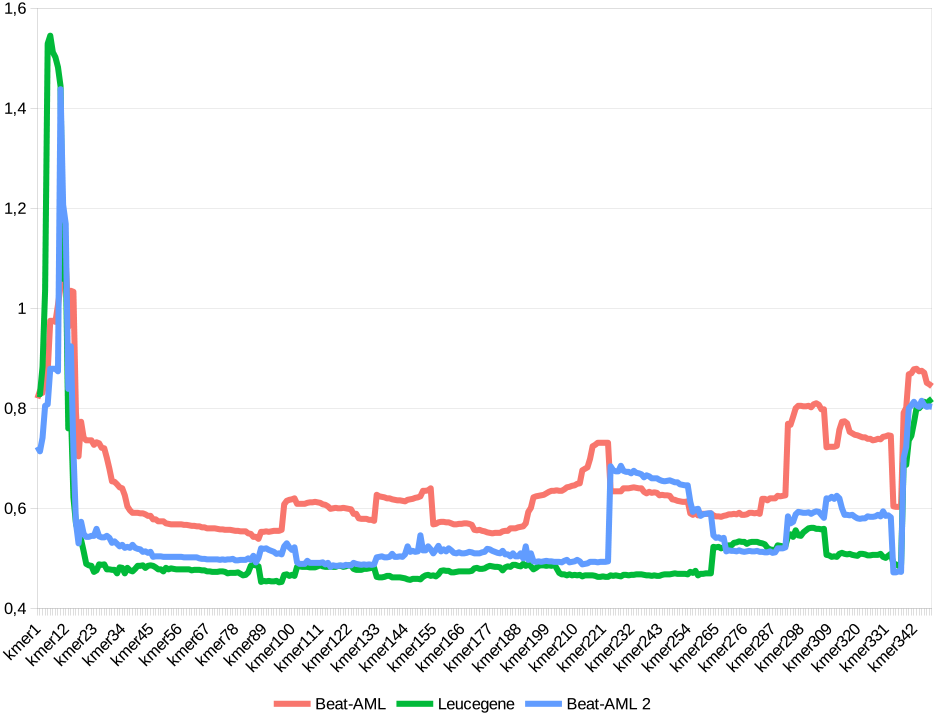
The coefficient of variation of k-mers in the AML cohorts for NPM1 gene.

In addition to the coefficient of variation between the conditions, to select the k-mer, we understood that it is interesting if, at least, half of one condition is different from zero, in this way, avoiding a k-mer containing a sample with an outlier expression.

### 2.3 Classifier models

To analyze if the selected k-mers are distinct between NPM1 mutated and non-mutated patients, we applied six of the most used Machine Learning algorithms to built the models and predict the conditions. For this task, considering that often the highest performing model are the least explainable and the most explainable are the least accurate (Gunning *et al*., 2021), we used two less complex models: K-nearest neighbors (KNN) and Logistic Regression (LR); and four complex models: Neural Network (NN), Random Forest (RF), Support Vector Machine (SVM), and eXtreme Gradient Boosting (XGB). We implemented the algorithms using Scikit-learn package (Pedregosa *et al*., 2011) in Python, applying for each model a grid search and a stratified cross-validation with 5-folds. We build models with normalized and non-normalized features. The k-mers count are considered the features, and the samples are the instances.

To understand how the k-mers can generalize between the cohorts, we constructed three settings to train and test the models, as shown in Figure 2, training on one cohort and testing on another cohort. The Setting A, where the models were trained on Beat-AML and tested on Leucegene. Setting B, where the models were trained on Leucegene and tested on Beat-AML2. Setting C, where the models were trained on Beat-AML and tested on Beat-AML2.

**Fig. 2:**
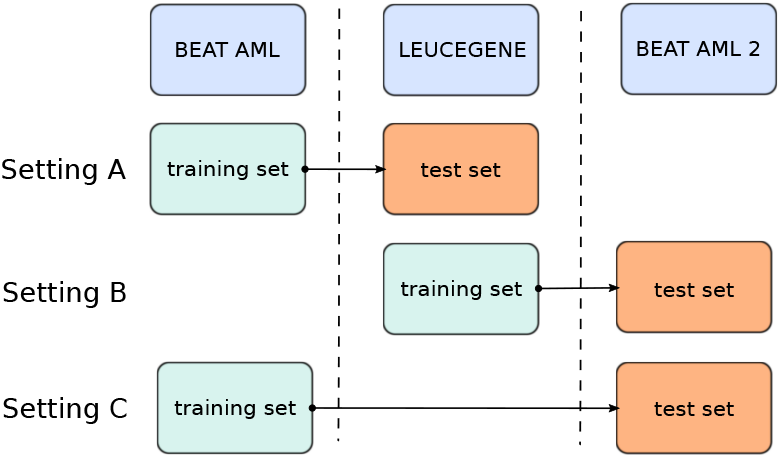
Schema to predict NPM1 mutated and non-mutated patients in cross-study scenarios.

The models were evaluated by area under the receiver operating characteristic (AUC) for each setting. Additionally, we evaluated the models by Accuracy, Kappa, F1-Score, Precision, and Recall metrics. Reproducible scripts can be found in https://osf.io/4s9tc/.

### 2.4 Mapping and Annotation

Our main objective was to identify the influence of the NPM1 mutation on k-mers and, consequently, on genes. To identify the genes belonging to the k-mers, we applied STAR 2.7.8a (Dobin and Gingeras, 2015) to map the k-mers to a reference human genome, GRCh38 assembly. After, we used Samtools 1.11 (Homer *et al*., 2009) to generate flexible alignment formats, SAM, BED, and BAM files.

Next, we implemented a script in R using the Ensembl REST API Yates *et al*., 2015 to request the genes annotation for each k-mer using the SAM and BAM files. Also, we generate BAM files for 27 samples: 9 NPM1 mutated and 9 NPM1 non-mutated (for every 9 samples, we selected 3 from Beat-AML, 3 from Leucegene, and 3 from Beat-AML 2), and 9 healthy samples (random selected). The 27 BAM files and the BED file from the k-mers were visualized using Integrative Genomics Viewer (IGV) version 2.13 (Robinson *et al*., 2011).

### 2.5 Model interpretation

An understanding of classifiers models is desirable when applying ML to health problems, however, most of the classifiers models are not easily explainable, requiring the use of Explainable Artificial Intelligence (XAI) tools. Seeking to get closer to the interpretation of the model and the selected k-mers, we applied the SHAP (Shapley Additive Explanations) tool (Lundberg and Lee, 2017) on themodelwith the best results to classify NPM1 mutated and non-mutated patients. SHAP is a tool to explain, by use of Shapley value (Winter, 2002), the prediction of each instance (sample) and the contribution of each feature (k-mer) to prediction. In addition, SHAP allows us to see the impact of these features across multiple instances, which means, showing the more important k-mers considering all the samples.

To see the important k-mers, and genes, we built a model with the intersection of k-mers between A, B, and C. We used only the annotated k-mers and the k-mers that do not belong to genes with high expression in the healthy cohort. After, we applied SHAP to the model. Reproducible scripts can be found in https://osf.io/4s9tc/.

## 3 Results

### 3.1 Model performance

As mentioned early, we constructed 5 classifier models to predict in three different settings. Figure 3 shows the performance of models using the AUC, in which, the XGB model had the best performance in the settings A, B, and C, with 97.3%, 94.2%, and 94.2%, respectively.

**Fig. 3:**
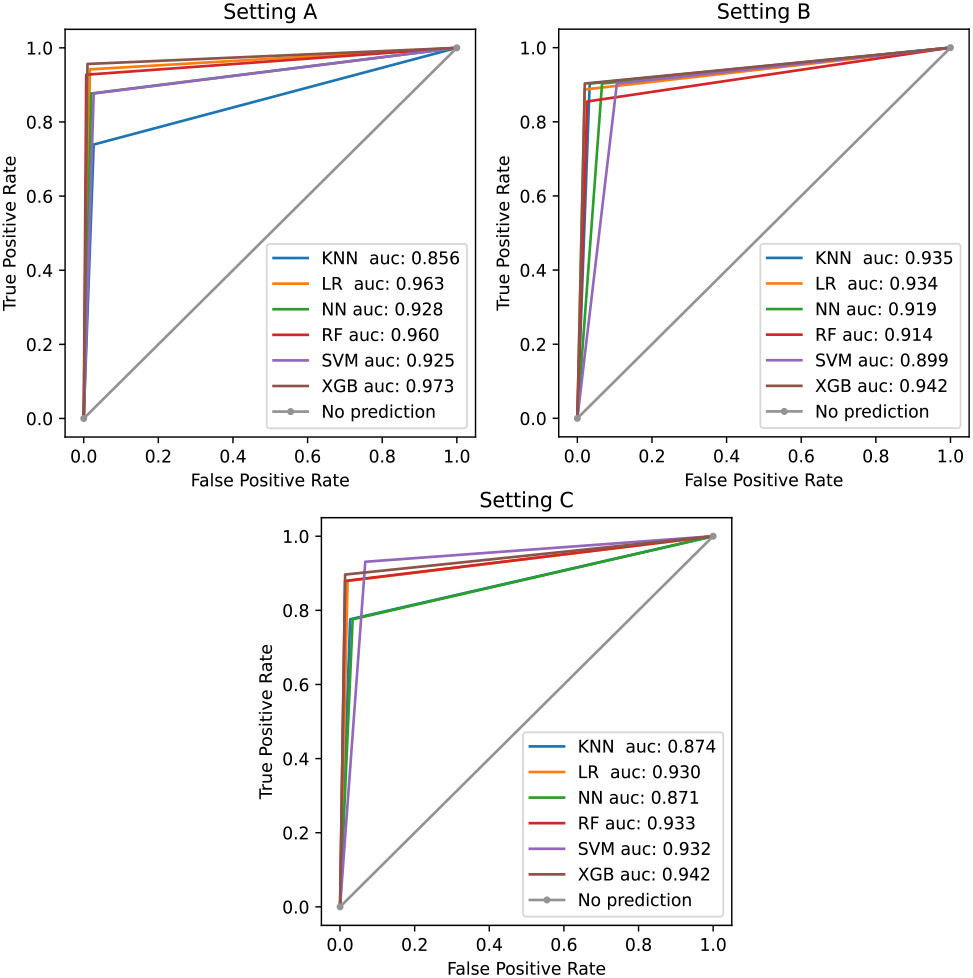
ROC curve and AUC metric applied to K-nearest neighbors (KNN), Logistic Regression (LR), Neural Network (NN), Random Forest (RF), Support Vector Machine (SVM), and eXtreme Gradient Boosting (XGB) models in the setting A, B, and C.

When analyzing the performance by Accuracy, Kappa, F1-Score, Precision, and Recall metrics, the XGB model achieves the best performance in all the metrics in setting A, as shown in Table 2.

**Table 2.** Accuracy, Kappa, F1-Score, Precision, and Recall metrics applied to K-nearest neighbors (KNN), Logistic Regression (LR), Neural Network (NN), Random Forest (RF), Support Vector Machine (SVM), and eXtreme Gradient Boosting (XGB) models in the setting A.

| Model | KNN | LR | NN | RF | SVM | XGB |
| --- | --- | --- | --- | --- | --- | --- |
| Accuracy | 0.899 | 0.970 | 0.947 | 0.972 | 0.942 | **0.979** |
| Kappa | 0.753 | 0.930 | 0.875 | 0.935 | 0.865 | **0.952** |
| F1-Score | 0.822 | 0.952 | 0.913 | 0.955 | 0.906 | **0.967** |
| Precision | 0.927 | 0.962 | 0.952 | 0.984 | 0.937 | **0.977** |
| Recall | 0.739 | 0.942 | 0.876 | 0.927 | 0.876 | **0.956** |

In setting B, the XGB model achieves the best performance in all the metrics. However, for the Recall metric, KNN, NN, and SVM have the same performance as XGB, as shown in Table 3.

**Table 3.**
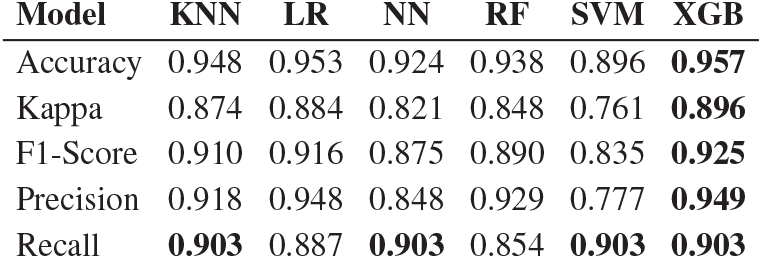
Accuracy, Kappa, F1-Score, Precision, and Recall metrics applied to K-nearest neighbors (KNN), Logistic Regression (LR), Neural Network (NN), Random Forest (RF), Support Vector Machine (SVM), and eXtreme Gradient Boosting (XGB) models in the setting B.

| Model | KNN | LR | NN | RF | SVM | XGB |
| --- | --- | --- | --- | --- | --- | --- |
| Accuracy | 0.948 | 0.953 | 0.924 | 0.938 | 0.896 | **0.957** |
| Kappa | 0.874 | 0.884 | 0.821 | 0.848 | 0.761 | **0.896** |
| F1-Score | 0.910 | 0.916 | 0.875 | 0.890 | 0.835 | **0.925** |
| Precision | 0.918 | 0.948 | 0.848 | 0.929 | 0.777 | **0.949** |
| Recall | **0.903** | 0.887 | **0.903** | 0.854 | **0.903** | **0.903** |

In setting C, the XGB model achieves the best performance in almost all the metrics, the exception was the Recall metric, where SVM has 93.1%. The XGB model has the second-best performance of Recall with 89.6%, as shown in Table 4.

**Table 4.** Accuracy, Kappa, F1-Score, Precision, and Recall metrics applied to K-nearest neighbors (KNN), Logistic Regression (LR), Neural Network (NN), Random Forest (RF), Support Vector Machine (SVM), and eXtreme Gradient Boosting (XGB) models in the setting C.

| Model | KNN | LR | NN | RF | SVM | XGB |
| --- | --- | --- | --- | --- | --- | --- |
| Accuracy | 0.917 | 0.951 | 0.912 | 0.956 | 0.931 | **0.960** |
| Kappa | 0.785 | 0.877 | 0.774 | 0.888 | 0.836 | **0.901** |
| F1-Score | 0.841 | 0.910 | 0.833 | 0.918 | 0.885 | **0.928** |
| Precision | 0.918 | 0.944 | 0.900 | 0.962 | 0.843 | **0.962** |
| Recall | 0.775 | 0.879 | 0.775 | 0.879 | **0.931** | 0.896 |

The majority of metrics were best performed by XGB models. The different results for the Recall metric can be explained due to the different proportions of imbalanced classes, once for imbalanced learning, Recall is typically used to measure the coverage of the minority class (Haibo and Yunqian, 2013). Between the three settings, setting B has the most balanced classes to train (31% to 68% in Leucegene), reflecting the learning of the minority class and being the easiest setting to measure by Recall, allowing KNN, NN, and SVM to perform well, as well as XGB.

On the other hand, setting C has the two most imbalanced cohorts to train (24% to 75% in Beat-AML) and test (28% to 71% in Beat-AML 2), making this the more difficult setting to measure by Recall. The good performance of SVM is due to the basic parameters of the SVM model, such as the C value, being able to treat the imbalance classes. In contrast, the XGB model needs specific parameters for imbalanced classes. The parameters and reproducible scripts for each model can be seen at https://osf.io/4s9tc/.

The results presented here belong to normalized models, performing better than non-normalized models. However, the results for non-normalized models only differ in 1% or 2% and can be found in the supplementary material.

### 3.2 Annotated genes

The selection step produced a different quantity of k-mers by setting, where the number of selected k-mers corresponded to the k-mers selected in the training cohort and found in the test cohort. For setting A, we selected 11,119 k-mers, of which 10,359 are completely aligned in the human genome, belonging to 93 annotated genes. For setting B, we selected 11,089 k-mers, of which 10,790 are completely aligned in the human genome, belonging to 58 annotated genes. For setting C, we selected 11,120 k-mers, of which 10,358 are completely aligned in the human genome, belonging to 93 annotated genes.

Figure 4 shows the number of genes by setting and the intersection between them. Setting A and C have the same 93 genes. The intersection genes between A and B, and B and C, are the same 30 genes. Thus, the intersection between the three settings is 30 genes.

**Fig. 4:**
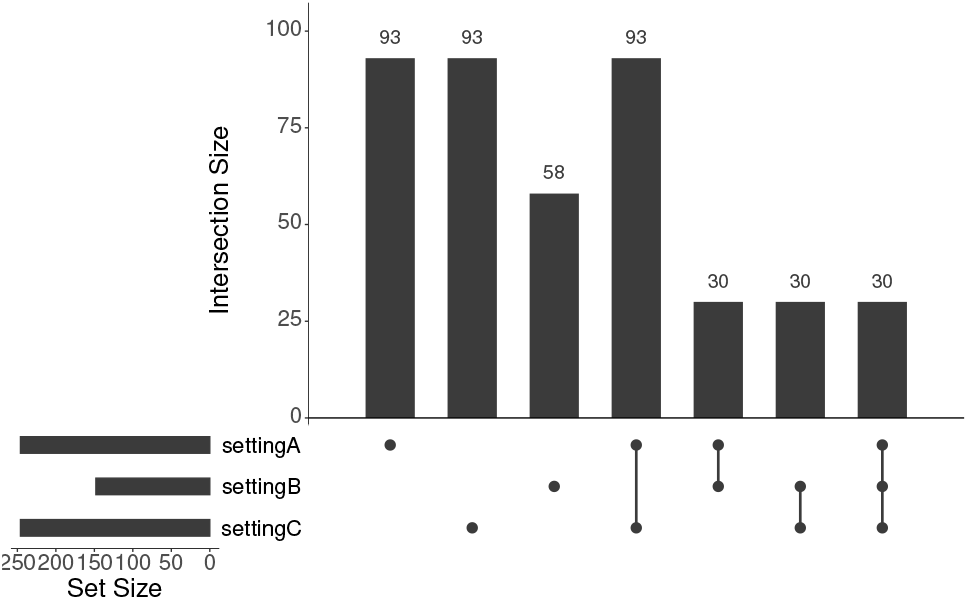
The number of genes annotated in setting A, B, and C. And intersections between them.

We analyzed the gene expression (based on k-mers average values) of the 30 intersection genes in the 3 AML cohorts (1112 samples), Figure 5. From 30 genes, 15 genes (in blue) show a different expression based on the statistical Wilcoxon test (comparison of average values) between NPM1 mutated and non-mutated patients. We presume that the other 15 genes influence the prediction indirectly, which means, the influence of these genes is not explained by the presence or absence of NPM1 mutation and needs to be studied in more detail.

**Fig. 5:**
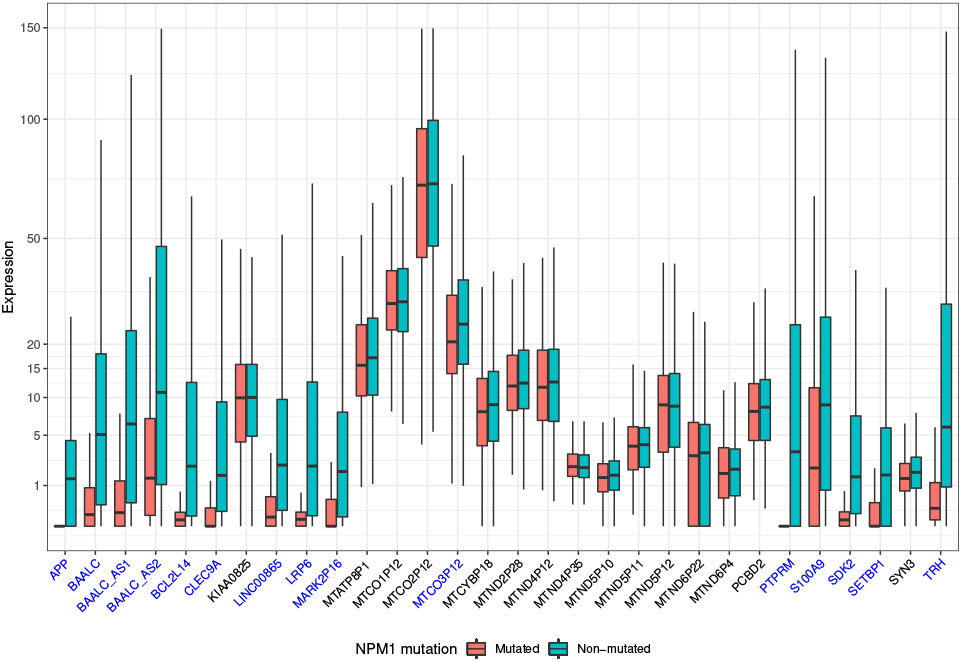
Gene expression (based on k-mers average values) of the intersection genes between the setting A, B, and C in the 3 AML cohorts. In blue, the genes with differential expression by statistical approach (comparison of average values by Wilcoxon test with Wilcoxon test P-Value less than 0.001) between NPM1 mutated and non-mutated patients.

Considering that we are looking for the genes directly impacted by NPM1 mutation for AML, we analyzed the 15 genes with differential expression in the healthy cohort, Figure 6. From 15 genes, 13 have gene expression (based on k-mers average values) less than 1. Even though we can not assume the suppression of these genes is linked with the presence or absence of NPM1 mutation, we can presume that these genes are linked with AML conditions.

**Fig. 6:**
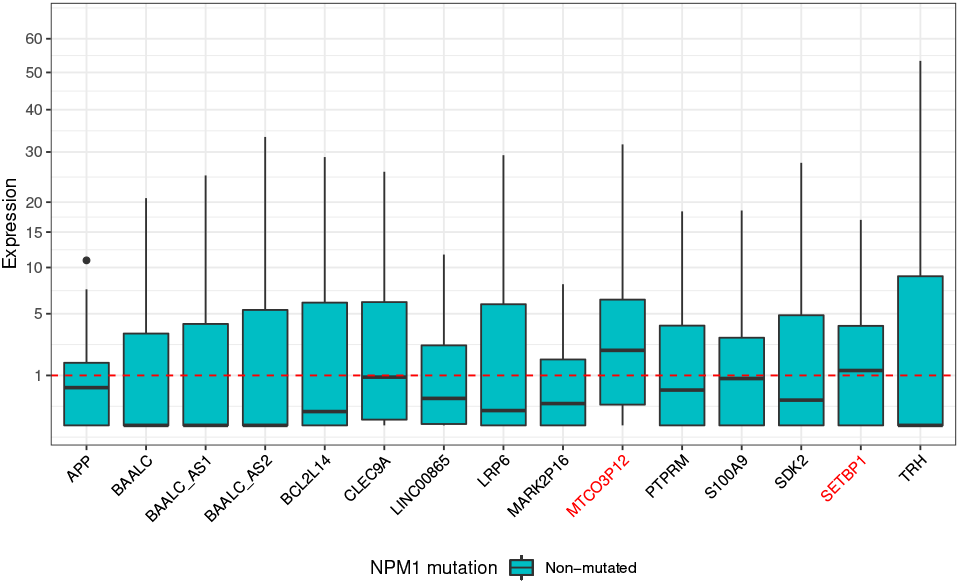
Gene expression (based on k-mers average values) of the intersection genes in the healthy cohort. In red, the genes with expression more than 1. The intersection genes that do not have differential expression between NPM1 mutated and non-mutated patients were removed in this analysis.

We then analyzed the gene expression of the 13 genes in a dataset of 34 samples corresponding to purified mature and immature blasts separated with specific surface markers from 17 AML patients. The expression analysis, Figure 7, shows that the 13 genes are expressed in the AML patients with a significant difference between mature and immature leukemia cells. Figure 7.A shows a tSNE (t-Distributed Stochastic Neighbour Embedding) analysis where the genes were able to group the mature and immature leukemia cells in different clusters. Figure 7.B presents that the genes are more expressed in immature leukemia cells. As AML cells with an immature phenotype play a central role in disease progression and relapse, the differential expression observed reinforces the involvement of these genes in AML.

**Fig. 7:**
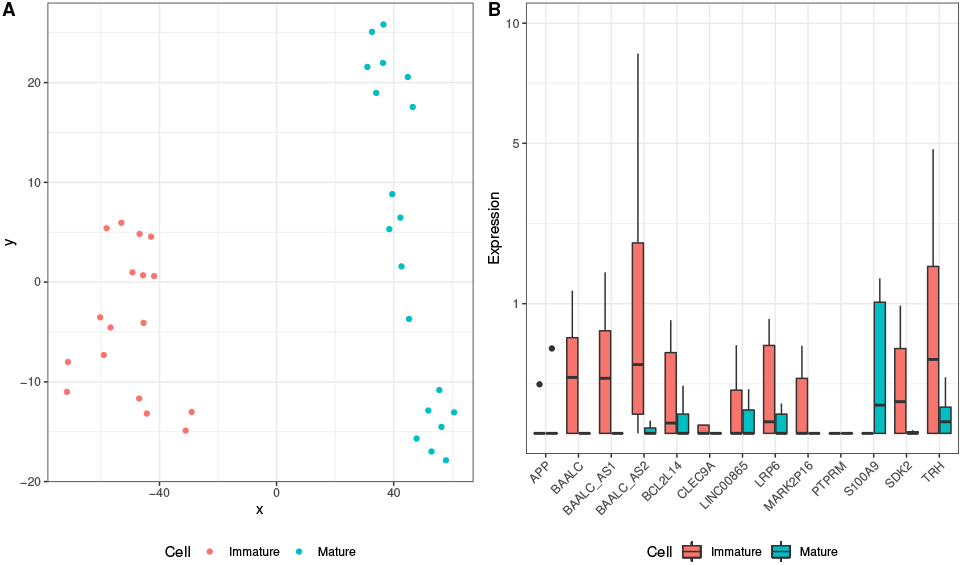
tSNE(A) and gene expression(B) analyses (based on k-mers average values) of the intersection genes in purified mature and immature blasts cells. The intersection genes that do not have differential expression between NPM1 mutated and non-mutated patients, and the genes with expression more than 1 in healthy cohort were removed in this analysis.

### 3.3 Model interpretation results

We constructed an XGB model using only the genes that the k-mers belonged to the 13 genes (we removed the k-mers from the genes without differential expression between the conditions and the genes with an expression of more than 1 in the healthy cohort). With this model, we used SHAP to identify the main k-mers, and genes, that impact the prediction of NPM1 mutation.

Figure 8 shows the 20 more important k-mers to the XGB model and their impact on prediction. On the left, we have the gene name belonging to each k-mer and the average SHAP value impact of the k-mer on the model. On the right, we have the impact of the k-mer on the model, where each point is a value for a k-mer and a patient. The colors are the values of the points (red for higher value, blue for lower value) and the X axis is the impact of SHAP value. Thus, analyzing the two more important k-mers, belonging to the SDK2 gene, the lower values of these k-mers have an impact on classification. However, regarding the third more important k-mer from the SDK2 gene, SHAP shows that the lower values of this k-mer have a negative impact.

**Fig. 8:**
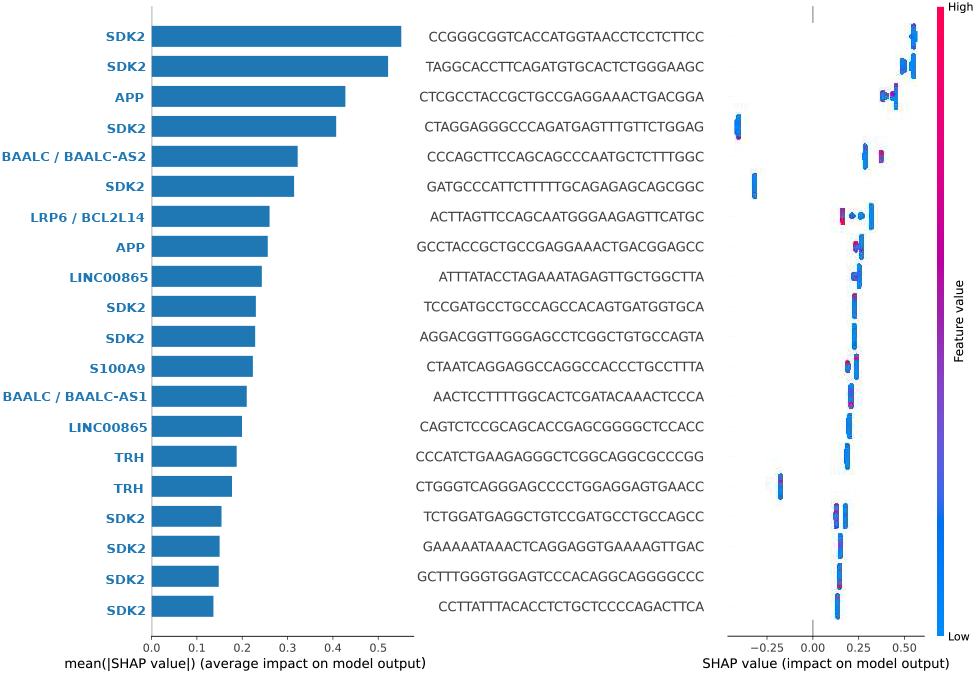
Top 20 more important k-mers, and the belonging genes, to XGB model by SHAP tool.

Once the best performance was achieved by one of the complex models, we tried to understand the k-mer selection by a more explainable model, the LR model. We build a LR model in the same conditions as the XGB model used by SHAP. After, we selected the more important k-mers by the coefficient of logistic regression, as showed in the Figure 9, presenting the coefficient weights for each k-mers, and the belonging genes.

**Fig. 9:**
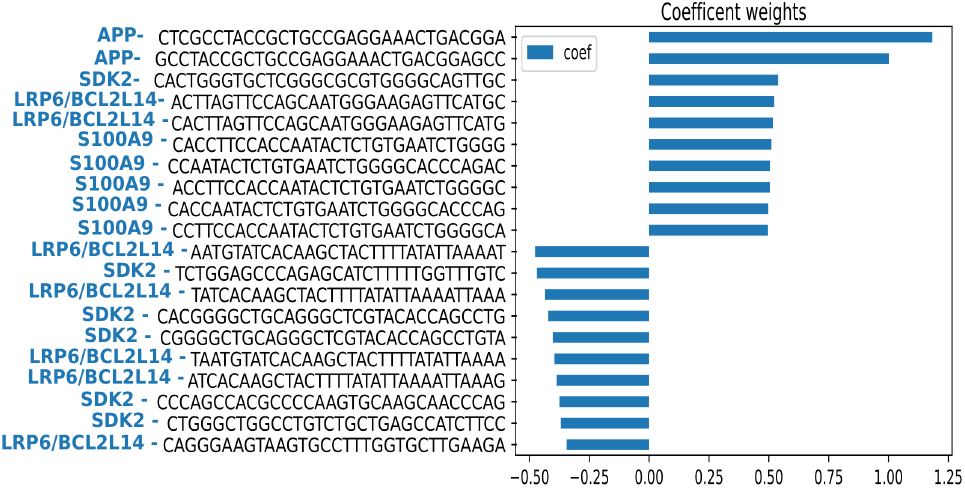
Top 20 more important k-mers, and the belonging genes, to LR model by coefficient of logistic regression.

SHAP interpreted that the more important k-mers to the XGB model belong to SDK2, APP, BAALC/BAALC-AS2, LRP6/BCL2L14, LINC00865, S1000A9, and TRH genes. LR model shows that the more important k-mers belong to APP, SDK2, LRP6/BCL2L14, and S100A9 genes. The two analyses have only the 3 k-mers in common, two belong to APP gene and one to LRP6/BCL2L14 gene. That shows us the need to further investigate the reasons for the choices made by the models and encourages us to understand the link between the most important k-mers.

### 3.4 Genes and k-mers expression

To get a better understanding of the selected k-mers at the gene level, we used IGV to visualize the distribution of the k-mers and reads in the groups of patients and healthy samples. We first compared the NPM1 expression and present a IGV view of 9 patients for each group (Figure 10). The NPM1 expression profile shows a read distribution similar between healthy, non-mutated, and mutated patients.

**Fig. 10:**
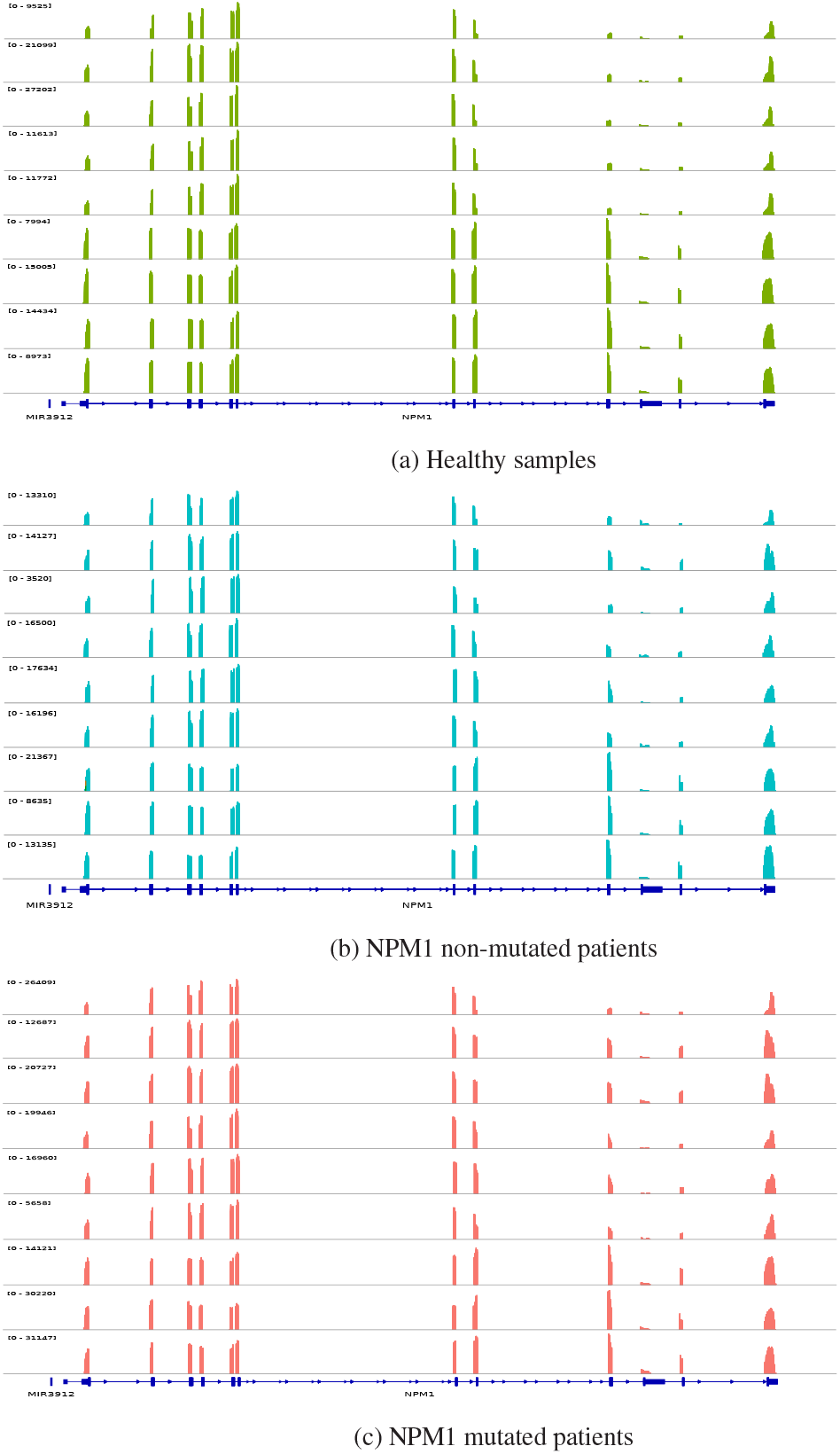
NPM1 gene expression at the gene level for healthy (a), non-mutated (b), and mutated (c) patients.

When analyzing the 13 genes from the selected k-mers, we observed a simple pattern for the TRH gene. As shown in Figure 11, the selected k-mers (the purple line at the bottom) cover almost the entire gene (the blue line at the bottom) at the exonic regions. Also, we observe a different profile in the TRH gene when comparing NPM1 mutated and non-mutated groups. Moreover, NPM1 non-mutated group presented the same profile as healthy donors.

**Fig. 11:**
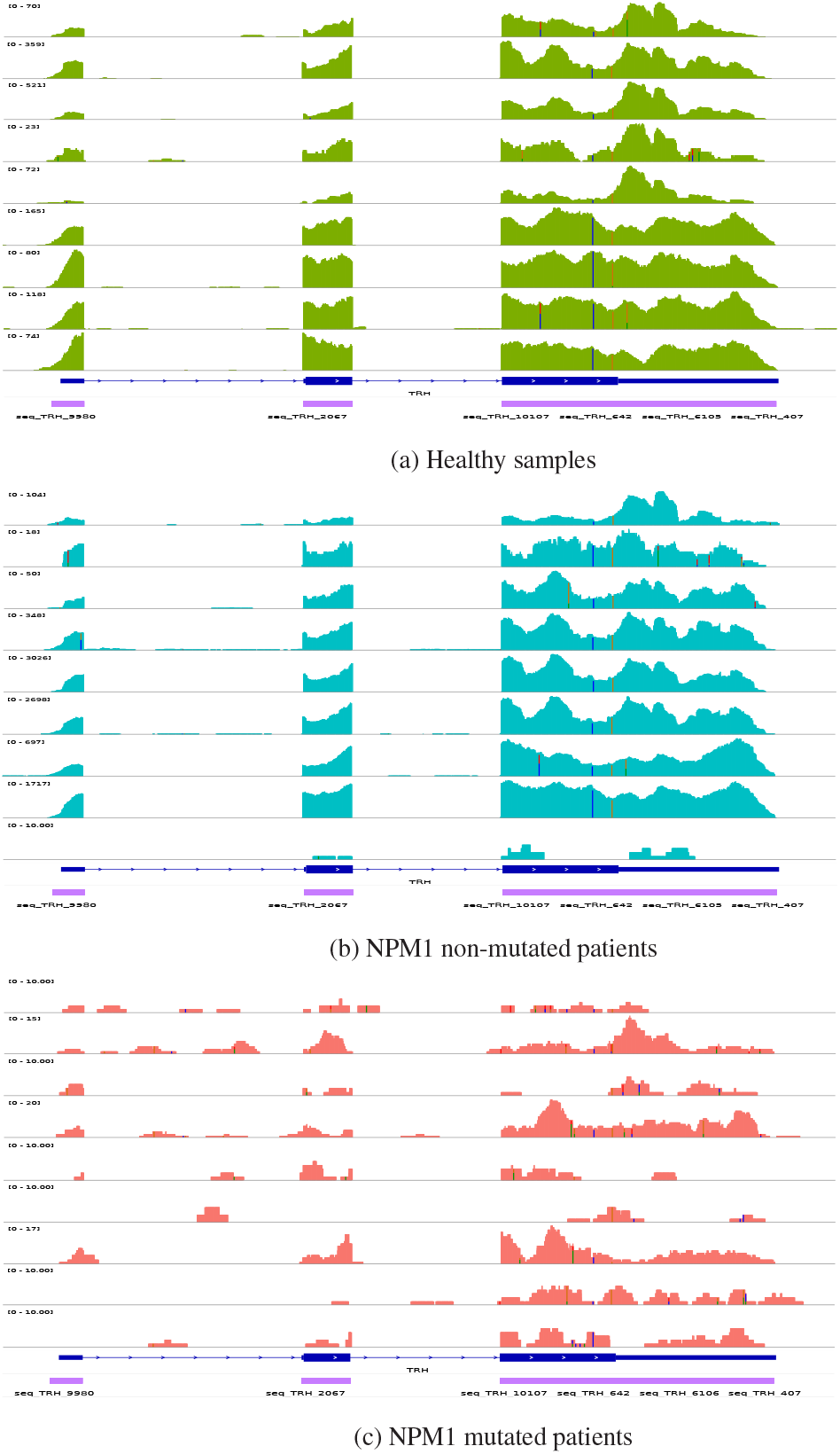
TRH gene expression at the gene level for healthy (a), non-mutated (b), and mutated (c) patients.

Investigating the APP gene, the gene with important k-mers in XGB and LR models, we could not find a simple pattern in the gene level as in the TRH gene. Thus, we investigated the APP gene at the k-mer level, as shown in Figure 12. We observed the same behavior, different profile for NPM1 mutated and non-mutated groups, and similar profile for NPM1 non-mutated and healthy groups. Also, at the k-mer level is possible to observe a low gene expression in NPM1 non-mutated patients.

**Fig. 12:**
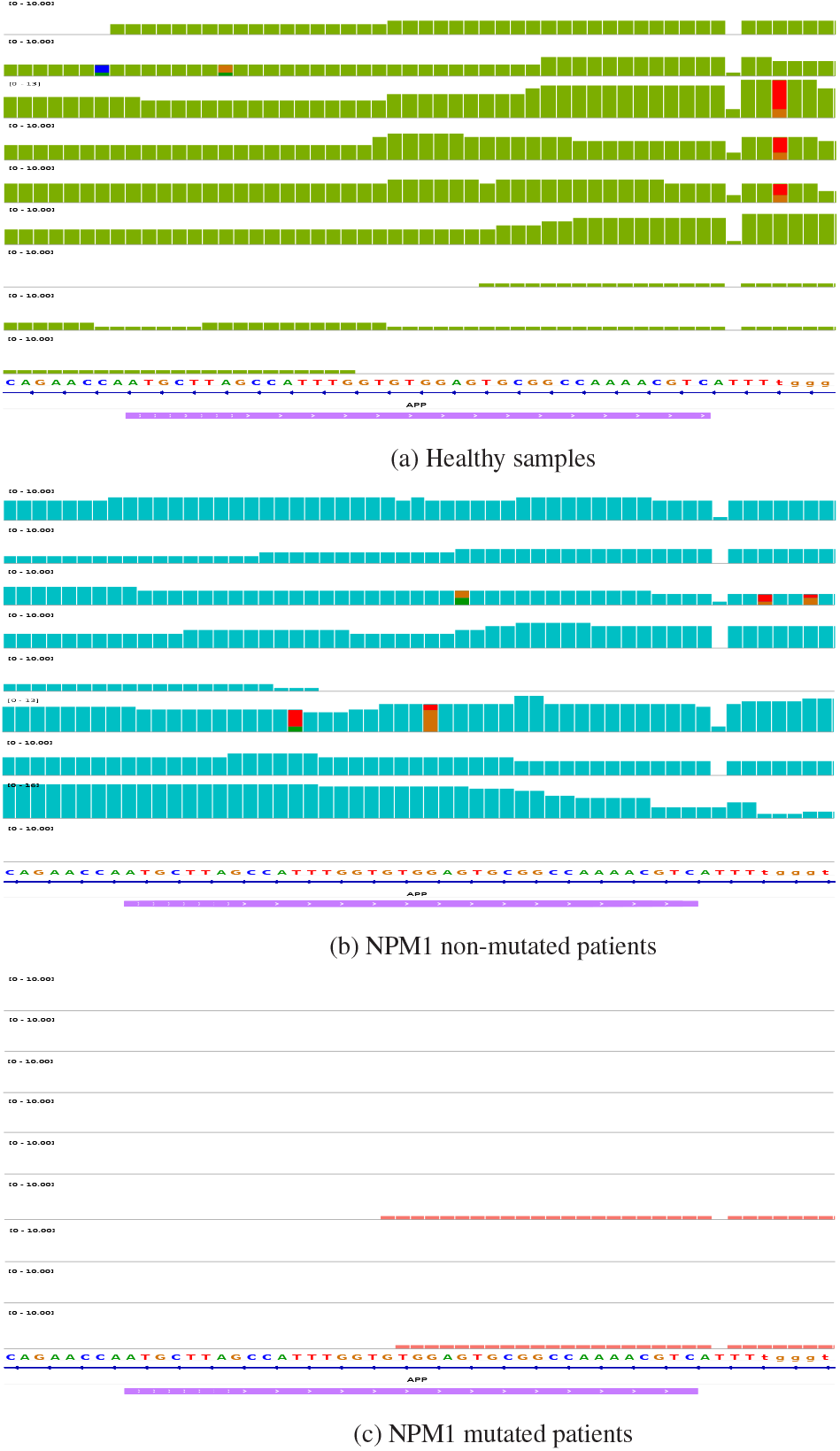
APP gene expression at the k-mer level for healthy (a), non-mutated (b), and mutated (c) patients.

Additionally, to understand the effect of NPM1 mutation in the APP gene, we analyzed the expression of NPM1 mutations and APP selected k-mers in Beat-AML cohort. The NPM1 mutations are at k-mer level (31nt), provided by Vizome, a platform with Beat-AML information. As showed in the Figure 13, in most cases, the patients that have a NPM1 mutation present low expression of APP selected k-mers as well as the patients that express the selected k-mers do not present the NPM1 mutation. That gives us a clear view of the interaction between the NPM1 mutation and APP at the k-mer level, interaction that we did not see at the gene level.

**Fig. 13:**
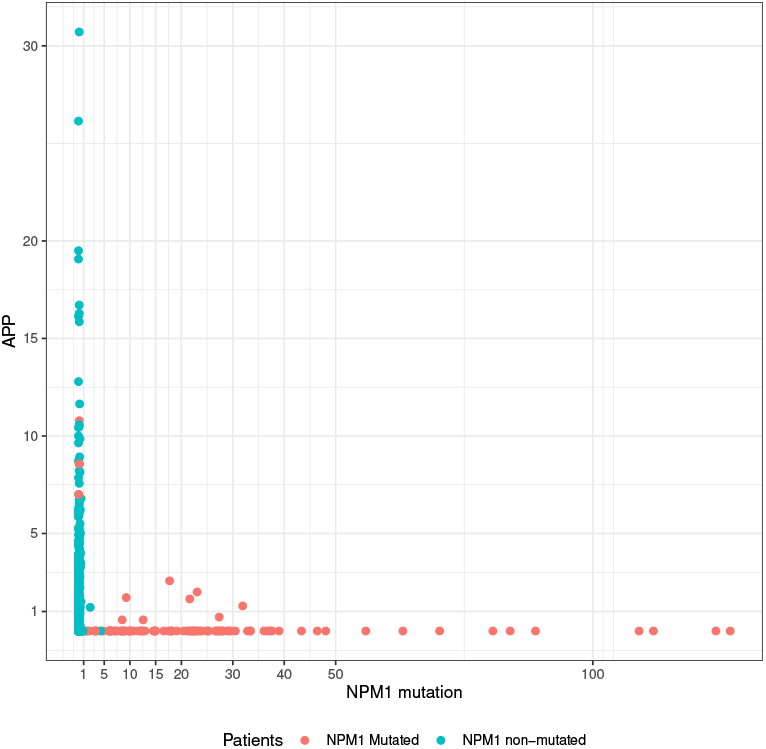
App and NPM1 mutation expression in Beat-AML patients.

## 4 Conclusion

Publicly available human RNA-seq datasets are precious resources for biomedical research and the analysis of existing datasets can be used to find a new information and search specific transcriptional events across patient cohorts. This required efficient tools and pipelines. As shown in our results, our pipeline, using k-mers, performed large datasets analyses corresponding to 1112 RNAseq samples. To predict and extract biological information, we constructed six classifier models with K-nearest neighbors (KNN), Logistic Regression (LR), Neural Network (NN), Random Forest (RF), Support Vector Machine (SVM), and eXtreme Gradient Boosting (XGB) algorithms applied on AML conditions based on the presence or absence of an NPM1 mutation. Comparing different settings built from the 3 cohorts, the XGB model achieves the best performance in the most metrics (AUC, Accuracy, Kappa, F1-Score, Precision, and Recall).

The pipeline works to find different gene expressions between conditions and the prediction models are able to be applied in different AML cohorts, once the models learned the diversity of the samples collected from bone marrow and peripheral blood, in different AML subtypes. Also, the pipeline can be further applied to other conditions defined by the presence of one or combined mutations described in AML groups, or to test new ones. Furthermore, the selected genes were not a random selection once some of them are already described in the literature as prognosis biomarkers or in link with NPM1 mutation deregulated pathways. Recently, Thyrotropin-Releasing Hormone (TRH) was identified as a novel AML prognosis biomarker downregulated in patients with NPM1 mutation (Gao *et al*., 2022). Alterations of BAALC expression are linked with molecular risk stratification (Prada-Arismendy *et al*., 2017) and have a possible linkage with a BCL2 family gene (Akhter *et al*., 2018). SETBP1 is an oncogene closely expressed in leukemia (Makishima, 2017). LRP6 and MARK2P16 belong to the WNT pathway required for leukemia stem cells development (Wang *et al*., 2010). Finally, CLEC9A is a lectin specifically expressed in immune cell subsets of the tumor microenvironment (Modak *et al*., 2022). Additionally, the genes found have different behaviors in AML and cohorts from healthy donors, showing specific deregulation in AML regardless of the NPM1 mutation condition.

As a perspective, the study requires an in-depth analysis of the selected genes, especially those with differential expression to find potential mechanisms of the NPM1 pathway from public chromatin and transcriptional regulation data (ReMap, JASPAR, Encode, FANTOM). Moreover, the k-mer level approach might be useful to detect complex transcriptional profiles, as shown for the APP gene, where the different expression can be seen only in the k-mer level. Thus, we will investigate more deply the dependence between the k-mers and to extend the analysis to search for differential events like splicing or intron retention.

## Data Availability

All data produced in the present study are available upon reasonable request to the authors.

https://github.com/railorena/npm1aml

