## supplementary material for "A k-mer based transcriptomics analysis for NPM1-mutated AML"

### Healthy Cohort

| Title/Cohort | Samples | Accession ID | Cell type |
|---|---|---|---|
| Functional Genomic Landscape of Acute Myeloid Leukemia | 33 | phs001657.v1.p1 | CD34+ and mononuclear |
| RNA-Seq analysis of human adult peripheral blood populations | 41 | GSE51984 | CD34+ and purified monocytes |
| Human Tumor-Associated Macrophage and Monocyte Transcriptional Landscapes Reveal Cancer-Specific Reprogramming, Biomarkers, and Therapeutic Targets | 22 | GSE117970 | purified monocytes |
| The Transcriptome Of Cmml Monocytes Is Highly Inflammatory And Reflects Leukemia-Specific And Age-Related Alterations | 28 | GSE135902 | monocytes |
| LncRNAs specific signature in acute myeloid leukemia with intermediate risk | 8 | GSE62852 | CD34+ |

**Parameters used in Kmtricks tools**

- Beat-AML

kmtricks pipeline --file samples --run-dir temp2 --kmer-size 31 -t 12 --hard-min 4 --soft-min 6 --recurrence-min 23 --focus 0 --mode kmer:count:bin --cpr --until merge
kmtricks aggregate --run-dir temp2 --matrix kmer --cpr-in --format text --sorted -t 12 > sorted_matrix.tsv

- Leucegene

kmtricks pipeline --file samples_leucegene --run-dir temp --kmer-size 31 -t 12 --hard-min 4 --soft-min 6 --recurrence-min 21 --focus 0 --mode kmer:count:bin --cpr --until merge
kmtricks aggregate --run-dir temp --matrix kmer --cpr-in --format text --sorted -t 12 > sorted_matrix.tsv

- Beat-AML 2

kmtricks pipeline --file samples_beat2 --run-dir temp --kmer-size 31 -t 12 --hard-min 4 --soft-min 6 --recurrence-min 10 --focus 0 --mode kmer:count:bin --cpr --until merge
kmtricks aggregate --run-dir temp --matrix kmer --cpr-in --format text --sorted -t 12 > sorted_matrix.tsv

- Healthy cohort

kmtricks pipeline --file samples --run-dir temp --kmer-size 31 -t 12 --hard-min 2 --focus 0 --mode kmer:count:bin --cpr --until merge
kmtricks aggregate --run-dir temp --matrix kmer --cpr-in --format text --sorted -t 12 > sorted_matrix.tsv

- Mature and immature blasts

kmtricks pipeline --file samples_scTCGA --run-dir temp --kmer-size 31 -t 12 --hard-min 2 --soft-min 4 --recurrence-min 5 --focus 0 --mode kmer:count:bin --cpr --until merge
kmtricks aggregate --run-dir temp --matrix kmer --cpr-in --format text --sorted -t 12 > sorted_matrix.tsv

### Models performance with non normalized and normalized models

- Setting A

| | Non normalized | | Normalized | |
|---|---|---|---|---|
| | **Accuracy** | **Kappa** | **Accuracy** | **Kappa** |
| **KNN** | 81.92 | 52.72 | 89.93 | 75.35 |
| **LR** | 87.41 | 67.93 | 97.02 | 93.07 |
| **NN** | 81.23 | 51.03 | 94.73 | 87.55 |
| **RF** | 96.56 | 91.88 | 97.25 | 93.54 |
| **SVM** | 79.86 | 55.97 | 94.27 | 86.52 |
| **XGB** | 96.79 | 92.61 | 97.94 | 95.20 |

- Setting B

| | Non normalized | | Normalized | |
|---|---|---|---|---|
| | **Accuracy** | **Kappa** | **Accuracy** | **Kappa** |
| **KNN** | 85.44 | 66.78 | 94.83 | 87.42 |
| **LR** | 90.61 | 77.03 | 95.30 | 88.40 |
| **NN** | 86.85 | 69.59 | 92.48 | 82.13 |
| **RF** | 95.30 | 88.40 | 93.89 | 84.85 |
| **SVM** | 84.03 | 57.25 | 89.67 | 76.10 |
| **XGB** | 94.36 | 86.34 | 95.77 | 89.61 |

- Setting C

| | Non normalized | | Normalized | |
|---|---|---|---|---|
| | **Accuracy** | **Kappa** | **Accuracy** | **Kappa** |
| **KNN** | 83.00 | 51.66 | 91.74 | 78.59 |
| **LR** | 87.86 | 67.44 | 95.14 | 87.74 |
| **NN** | 82.52 | 52.30 | 91.26 | 77.45 |
| **RF** | 94.17 | 84.97 | 95.63 | 88.91 |
| **SVM** | 85.43 | 65.10 | 93.20 | 83.71 |
| **XGB** | 96.11 | 90.19 | 96.11 | 90.19 |
